# The contribution of sub-optimal prescription of preoperative antiplatelets and statins to race and ethnicity-related disparities in major limb amputation

**DOI:** 10.1101/2023.02.20.23286203

**Authors:** Corey A. Kalbaugh, Brian Witrick, Kerry A. Howard, Laksika Banu Sivaraj, Katharine L. McGinigle, Samuel Cykert, William P. Robinson, Catherine R. Lesko

## Abstract

**Background:** People undergoing revascularization for symptomatic peripheral artery disease (PAD) have a high incidence of major limb amputation in the year following their surgical procedure. The incidence of limb amputation is particularly high in patients from racial and ethnic minority groups. The purpose of our study was to investigate the role of sub-optimal prescription of preoperative antiplatelets and statins in producing disparities in risk of major amputation following revascularization for symptomatic PAD.

**Methods:** We used data from adult (≥18 years old) patients in the Vascular Quality Initiative (VQI) registry who underwent a revascularization procedure from 2011-2018. Patients were categorized as non-Hispanic Black, non-Hispanic White, and Hispanic. We estimated the crude probability of a patient being prescribed a preoperative antiplatelet and preoperative statin. We calculated one year risk incidence of amputation by prescription groups and by race/ethnicity. We estimated the amputation risk difference between race/ethnicity groups (the proportion of disparity) that could be eliminated under a hypothetical intervention where a pre-operative antiplatelet and statin was provided to all patients.

**Results:** Across 100,579 revascularizations recorded in the Vascular Quality Initiative, a vascular procedure-based registry in the United States and Canada, 1-year risk of amputation was 2.5% (95% CI: 2.4%,2.6%) in White patients, 5.3% (4.9%,5.6%) in Black patients and 5.3% (4.7%,5.9%) in Hispanic patients. Black (57.5%) and Hispanic patients (58.7%) were only slightly less likely than White patients (60.9%) to receive recommended antiplatelet and statin therapy prior to their procedures. However, the effect of antiplatelets and statins was greater in Black and Hispanic patients such that, had all patients received the appropriate guideline recommended medications, the estimated risk difference comparing Black to White patients would have reduced by 8.9% (−2.9%,21.9%) and the risk difference comparing Hispanic to White patients would have been reduced by 17.6% (−0.7%,38.6%).

**Conclusions:** Even though guideline-based care appeared evenly distributed by race/ethnicity, increasing access to such care may still decrease health care disparities in major limb amputation.

## INTRODUCTION

Peripheral artery disease (PAD) is a common atherosclerotic disorder that reduces blood flow to the lower extremities, leading to muscle pain, ischemic wounds that do not heal, and eventual tissue death^1^. Globally, more than 200 million people have PAD with a disproportionate number living in low and middle income countries^2^. Among the 8.5-12 million people estimated to have PAD in the United States^3^, the highest burden and worst prognosis following a PAD diagnosis are consistently observed in historically disadvantaged groups such as Black and Hispanic-Latin-X (hereafter referred to as Hispanic) populations^4, 5^.

Black people in the United States have a higher prevalence and incidence of PAD,^6^ receive less preventive treatment including for secondary prevention,^7^ and have worse limb-based outcomes^8^ compared to white people. In addition, white patients with PAD are more likely than black patients to have a revascularization (i.e. a limb-sparing procedure) attempt for their PAD rather than an amputation (i.e. limb-removing procedure)^9^. Black patients with symptomatic PAD are 4-5 times more likely than white patients to undergo a PAD-related limb amputation^10, 11^. While the incidence of PAD in Hispanic versus non-Hispanic white people may be similar^12, 13^, Hispanic people with PAD are under-prescribed cardiovascular medications^14^ and have a higher incidence of chronic limb-threatening ischemia (CLTI; end-stage PAD) when hospitalized^5^. Hispanic patients are also more likely to receive a limb amputation compared to non-Hispanic white patients^15, 16^.

Consistency of care remains an important topic in the vascular community where guidelines are written – and updated regularly – to optimize care^17, 18^. However, little is known about the uptake of PAD guidelines, particularly in diverse populations. We do know that when PAD guidelines are followed, outcomes for patients with PAD are improved^19-21^. As one potential explanation of the poor limb outcomes observed in Black and Hispanic patients, we hypothesized that guideline-directed pre-operative management may be differentially administered across race/ethnicity groups and that differentially applied care may increase harm to already disadvantaged race/ethnicity groups.

Herein, we focused on one component of guideline-directed preoperative care for patients undergoing revascularization for PAD: the prescription of antiplatelets and statins prior to a revascularization procedure. We described the prevalence of prescriptions of antiplatelets and statins; estimated the effect of prescribing antiplatelets and statins on subsequent risk of amputation stratified by race/ethnicity; and estimated the proportion of race/ethnicity-related disparities in major limb amputation following vascular intervention for PAD that could be eliminated through universal provision of pre-operative antiplatelets and statins.

## METHODS

### Database

The Society for Vascular Surgery (SVS) Patient Safety Organization established the Vascular Quality Initiative (VQI), a vascular procedure-based registry to document and improve the quality of care delivered to patients with vascular diseases^22^. Clinicians that participate in the delivery of vascular care in the United States or Canada are eligible to participate in the VQI. A map of participating sites is included in the supplemental files (Figure S1). During our study period, 5,215 physicians at 536 healthcare centers were participating in a VQI registry. Centers include academic medical centers, teaching hospitals, community hospitals, and private practices.

A detailed description of the VQI registry has been previously published^22^. Briefly, trained data abstractors or clinicians input all data from each Center into one of fourteen major vascular procedure registries. Each registry contains information on patient demographic traits, comorbid conditions, imaging studies, medication usage, peri-procedural details, and in-hospital and thirty-day outcomes. Patients also complete a one-year follow-up visit for any procedure covered by the VQI and outcome data are included in the registry.

### Study sample

This study used data from adult (≥18 years old) patients in the VQI registry who underwent a revascularization procedure from 2011-2018. We restricted our analysis to revascularization procedures indicated due to chronic PAD including claudication (pain while walking) or CLTI (inadequate blood and oxygen supply resulting in ischemic pain at rest or tissue loss). The registries also capture patients treated for acute ischemia (15%), a different clinical etiology that is excluded from our analysis. The Institutional Review Board of Clemson University determined the study met criteria for exemption because only deidentified data were used.

The unit of analysis for this study was an affected revascularized leg; individual patients could contribute two legs to this analysis. We included only the first procedure for the right and left legs of each patient. Patients of race-ethnicity other than non-Hispanic Black, non-Hispanic White, and Hispanic were also excluded from analyses. Hispanic ethnicity was defined as a person of Cuban, Mexican, Puerto Rican, South or Central American, or other Spanish culture or origin, regardless of race.

### Outcome

Our outcome was major amputation, defined as an amputation that occurred either below the knee or above the knee but excluding minor amputations of toes or trans-metatarsal or midfoot. Major amputation is most often considered a failure of vascular care and is associated with loss of mobility and independent living status,^23, 24^ as well as increased risk of subsequent mortality^25^.

### Guideline Care Measures

The major organizations that govern vascular care, including the American College of Cardiology and the American Heart Association (ACC/AHA) and the SVS, recommend that patients with PAD receive antiplatelet therapy and statin medications for secondary prevention and cardiovascular risk reduction^17, 18^. Patients who took aspirin, or drugs that include aspirin, within 36 hours prior to surgery were classified as following antiplatelet therapy recommendations. Patients who received any of the HMG-CoA reductase inhibitors used to reduce cholesterol and stabilize atherosclerotic plaque within 36 hours prior to surgery were classified as following the recommendations for statin medications.

### Demographics and Comorbidities

All demographics and comorbidities were abstracted from the medical record at each participating center. Demographics included race, ethnicity, sex, and age at time of procedure (modeled as a quadratic). Insurance status was determined by the primary payer of the procedure and classified as uninsured (none, self-pay) or insured (Medicare, Medicaid, Commercial, Military/VA, non-US insurance). Smoking history was categorized as never or ever. Diabetes mellitus was defined as absent or present (whether controlled or uncontrolled). Chronic obstructive pulmonary disease (COPD) was categorized as absent, present and requires medication, or present and requires home oxygen. Hypertension was defined as documented history of blood pressure ≥ 140 mm Hg/90 mm Hg on at least 3 occasions. Obesity was defined as having a body mass index ≥ 30 kg/m^2^. Coronary artery disease was classified as present if there was any history of myocardial infarction, stable angina, or unstable angina. Congestive heart failure was defined according to the New York Heart Association Classification system^26^ and classified as present if patients were asymptomatic with a positive history or symptomatic regardless of severity. Dialysis-dependence was defined as current use of hemo-or peritoneal dialysis. We categorized disease severity (indication for revascularization) as claudication or CLTI. Revascularization was classified as either 1) infrainguinal peripheral vascular intervention (PVI) including percutaneous catheter-based revascularization or 2) infrainguinal bypass.

### Statistical analysis

We first compared the distribution of demographic characteristics and comorbidities of patients in our study sample across race/ethnicity groups. We then compared the crude probability of having a preoperative antiplatelet, a preoperative statin, and both antiplatelets and a statin, across race/ethnicity groups, in the entire sample and stratified by the indication for revascularization (CLTI or claudication).

We followed patients/limbs from revascularization until the first occurrence of major amputation, death (a competing event), or administrative censoring at one year. We report results based on the 30-day and 1-year cumulative incidence of major amputation, estimated using the Aalen-Johansen estimator^27^. For all analyses, we accounted for possible selection bias as a result of differential loss to follow-up with inverse probability of censoring weights conditional on all baseline covariates defined above and weeks since procedure^28^.

To estimate whether the effect of guideline-directed provision of antiplatelets and statins differed by race/ethnicity, we estimated risk differences (RD) for having been prescribed both a preoperative antiplatelets and statin compared to only one or neither medication, stratified by race/ethnicity.

To estimate the impact of universal provision of pre-operative antiplatelets and statins on racial/ethnic disparities, we used the framework described in Howe, et al^29^. We first estimated the total association (“ total effect,” TE, although we do not interpret this association causally) between race/ethnicity and 30-day and 1-year cumulative incidence of major amputation comparing non-Hispanic Black and Hispanic patients with non-Hispanic white patients (the referent group). These estimates were only adjusted for possible selection bias with inverse probability of censoring weights.

We next estimated the 30-day and 1-year RDs associated with race/ethnicity *that we would expect to see* if all patients had been prescribed pre-operative antiplatelets and statins (the “ controlled direct effect,” CDE, although again, we do not interpret this association causally). We estimated the cumulative incidence of amputation following revascularization procedures restricted to patients who received both a pre-operative antiplatelets and statin, again stratified by race/ethnicity. We accounted for differences in who did and did not receive guideline-directed management using inverse probability of the mediator weights. We estimated these weights from generalized logistic models for the probability that both antiplatelet and statin were prescribed preoperatively, relative to one drug only, or neither drug, conditional on race/ethnicity and all covariates described above. We stabilized these weights on race/ethnicity.

The proportion of the racial/ethnic disparity eliminated (PE) under a hypothetical intervention to ensure a pre-operative antiplatelet and statin was provided to all patients was estimated as (TE-CDE)/TE.

To account for uncertainty in the weights, and also possible correlation between major amputations for limbs belonging to the same individual, we estimated 95% confidence intervals for all estimates described above by selecting the 2.5^th^ and 97.5^th^ percentiles from the distribution of such estimates from 500 bootstrap resamples of the data. Bootstrap resamples were conducted through the Palmetto Cluster for high performance computing at Clemson University. All analyses were performed using SAS version 9.4 (SAS Institute Inc., Cary, NC).

### Secondary Analyses

We also estimated the PE in subpopulations of revascularization procedures that were indicated for claudication and CLTI.

## RESULTS

### Study Population

We identified 100,579 revascularizations (limbs) among 88,599 patients within the VQI. Median follow-up time was 365 days (84% of limbs were followed for a full year). The population was comprised of mostly male patients (63%), and mostly Non-Hispanic White patients (76%). There were 15,442 Non-Hispanic Black patients (17%), and 5,506 Hispanic patients (7%). The majority of revascularizations (57%) were for an indication of CLTI. More than 50% of patients had diabetes (54%) and hypertension (89%) prior to revascularization. These conditions were more prevalent in Black and Hispanic patients than Non-Hispanic White patients, as was dialysis dependency, while White patients were more likely to have COPD (Table 1).

**Table I.** Characteristics (number and percent) of patients^a^ who underwent revascularization for peripheral arterial disease with an indication of chronic limb-threatening ischemia or claudication, captured by the Vascular Quality Initiative registry, United States, 2011-2018, stratified by race/ethnicity group

|  | **Overall** | **Non-Hispanic White** | **Non-Hispanic Black** | **Hispanic** |
| --- | --- | --- | --- | --- |
| **N** | **88,599** | **67,651** | **15,442** | **5,506** |
| **Characteristics of the procedure** |  |  |  |  |
| Bypass | 23,556 (26.6) | 18,204 (26.9) | 3,887 (25.2) | 1,465 (26.6) |
| Disease subtype: CLTI | 50,250 (56.7) | 36,395 (53.8) | 10,145 (65.7) | 3,710 (67.4) |
| **Patient characteristics** |  |  |  |  |
| Mean (SD) Age, years | 68.6 (11.2) | 69.3 (11.0) | 65.8 (11.2) | 67.8 (11.6) |
| Female sex | 32,878 (37.1) | 23,675 (35.0) | 7,048 (45.6) | 2,155 (39.2) |
| Current smoker | 29,759 (33.6) | 22,877 (33.8) | 5,613 (36.4) | 1,269 (23.1) |
| Diabetes | 47,628 (53.8) | 33,930 (50.2) | 9,624 (62.3) | 4,074 (74.0) |
| Obese | 29,550 (33.5) | 22,592 (33.6) | 5,305 (34.5) | 1,653 (30.2) |
| Hypertension | 78,704 (88.9) | 59,387 (87.8) | 14,327 (92.8) | 4,990 (90.7) |
| Coronary artery disease | 28,316 (32.0) | 22,492 (33.3) | 4,121 (26.7) | 1,703 (30.9) |
| Congestive heart failure | 16,688 (18.8) | 12,335 (18.2) | 3,307 (21.4) | 1,046 (19.0) |
| COPD | 21,944 (24.8) | 18,066 (26.7) | 3,021 (19.6) | 857 (15.6) |
| Dialysis | 7,533 (8.5) | 3,941 (5.8) | 2,690 (17.4) | 902 (16.4) |

Abbreviations: SD=standard deviation; CLTI=chronic limb-threatening ischemia; COPD=chronic obstructive pulmonary disease
<sup>a</sup> Patients could contribute up to 2 legs to the analysis; characteristics are presented for the first revascularization procedure in the registry

### Prescription Patterns

Patients received preoperative antiplatelet and statin medications before 60.2% of revascularizations. The proportion of revascularizations with a preoperative antiplatelet and statin was slightly higher for non-Hispanic White patients (60.9%) than for non-Hispanic Black patients (57.5%) and Hispanic patients (58.7%). These patterns held true across disease subtypes of CLTI and claudication (Table II).

**Table II.** Presence of guideline-recommended medication prescriptions within 36 hours prior to revascularization, by race-ethnicity and disease subtype, for n=100,579 revascularizations for chronic limb-threatening ischemia or claudication, captured by the Vascular Quality Initiative registry, United States, 2011-2018

| Population | Medication | Overall | Non-Hispanic White | Non-Hispanic Black | Hispanic | P-Value |
| --- | --- | --- | --- | --- | --- | --- |
| Overall |  |  |  |  |  |  |
|  | Antiplatelet | 75839 (75.4) | 58520 (76.1) | 12781 (73.3) | 4538 (73.6) | <0.0001 |
|  | Statin | 74579 (74.2) | 57527 (74.8) | 12498 (71.7) | 4554 (73.8) | <0.0001 |
|  | Antiplatelet + Statin | 60481 (60.2) | 46832 (60.9) | 10028 (57.5) | 3621 (58.7) | <0.0001 |
| CLTI |  |  |  |  |  |  |
|  | Antiplatelet | 40186 (72.5) | 29335 (73.1) | 7923 (70.7) | 2928 (71.5) | <0.0001 |
|  | Statin | 39599 (71.5) | 28870 (72.0) | 7806 (69.6) | 2923 (71.3) | <0.0001 |
|  | Antiplatelet + Statin | 31240 (56.4) | 22841 (56.9) | 6108 (54.5) | 2291 (55.9) | <0.0001 |
| Claudication |  |  |  |  |  |  |
|  | Antiplatelet | 35653 (79.0) | 29185 (79.3) | 4858 (77.9) | 1610 (77.7) | 0.0139 |
|  | Statin | 34980 (77.6) | 28657 (77.9) | 4692 (75.3) | 1631 (78.8) | <0.0001 |
|  | Antiplatelet + Statin | 29241 (64.8) | 23991 (65.2) | 3920 (62.9) | 1330 (64.2) | 0.0016 |

Abbreviation: CLTI=chronic limb-threatening ischemia.

### Impact of Preoperative Antiplatelet and Statin

In comparing amputation for those receiving preoperative prescription of antiplatelet + statin versus neither across race/ethnicity groups, the risk of amputation at one year was reduced among all groups. However, the reduction in amputation risk was more pronounced in Black (RD: 2.2%; 95% CI: 0.7, 3.8) and Hispanic (RD: 1.9%; 95% CI: -0.3, 4.2) patients compared to White (RD: 0.5%; 95% CI: 0.0, 0.9) patients (Table III).

**Table III.**
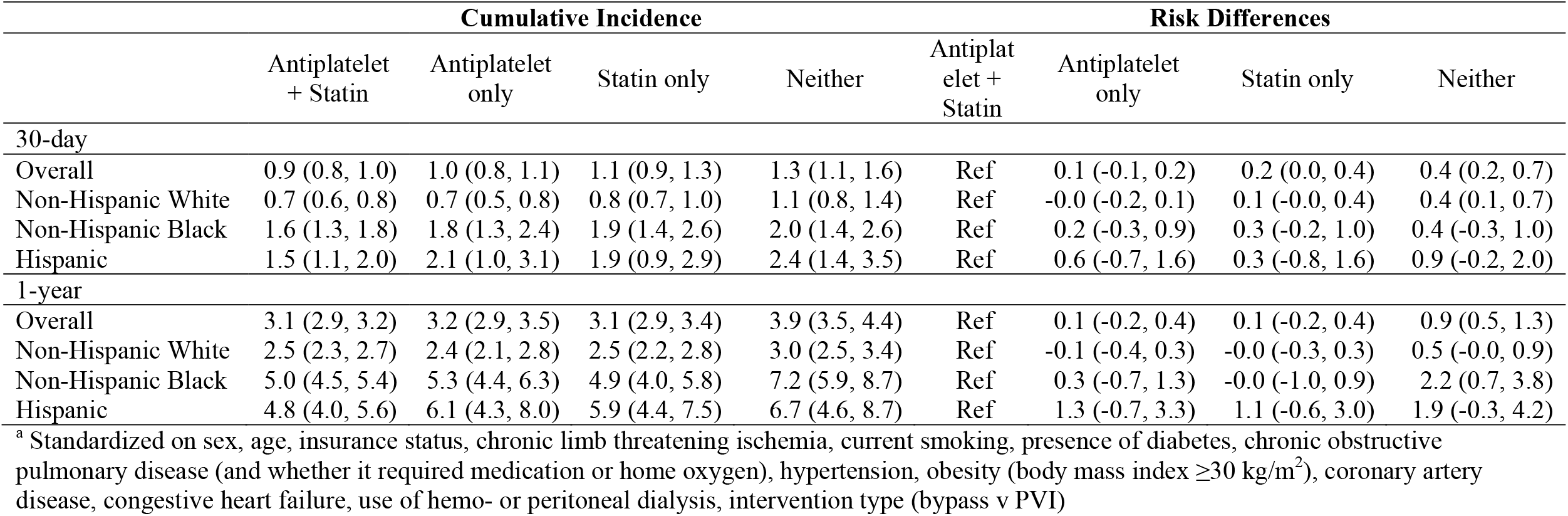
Standardized^a^ cumulative incidence and risk differences for major amputation associated with preoperative (within 36 hours) antiplatelet and statin non-use versus use for n=100,579 revascularizations for chronic limb-threatening ischemia or claudication, captured by the Vascular Quality Initiative registry, United States, 2011-2018

### Impact of Antiplatelet and Statins on Racial Disparities

The cumulative incidence of limb amputation among non-Hispanic White patients was 0.8% (95% CI: 0.7, 0.8) at thirty days and 2.5% (95% CI: 2.4, 2.6) at one year (adjusted only for loss to follow-up). Non-Hispanic Black patients and Hispanic patients had higher cumulative incidence rates of amputation than non-Hispanic White patients at thirty days, 1.7% (95% CI:1.5,1.9) and 1.8% (95% CI:1.5,2.1), respectively; and at one year, 5.3% (95% CI: 4.9,5.6) and 5.3% (95% CI:4.7,5.9), respectively.

Had all patients received the appropriate preoperative medication, the amputation risk would have been lower across all three race/ethnicity groups at both thirty days and one year (Table III). Compared to non-Hispanic White patients, the proportion of disparities that could be eliminated (i.e. the percent reduction in the risk difference observed under no intervention) for non-Hispanic Black patients was 8.6% (95% CI: -13.6, 28.8) at thirty days and by 8.9% (95% CI: -2.9, 21.9) at one year. For Hispanic patients, the proportion of disparities eliminated compared to non-Hispanic White patients was 17.2% (95% CI: -11.9, 50.1) at thirty days and 17.6% (95% CI: -0.7, 38.6) at one year (Table IV).

**Table IV.**
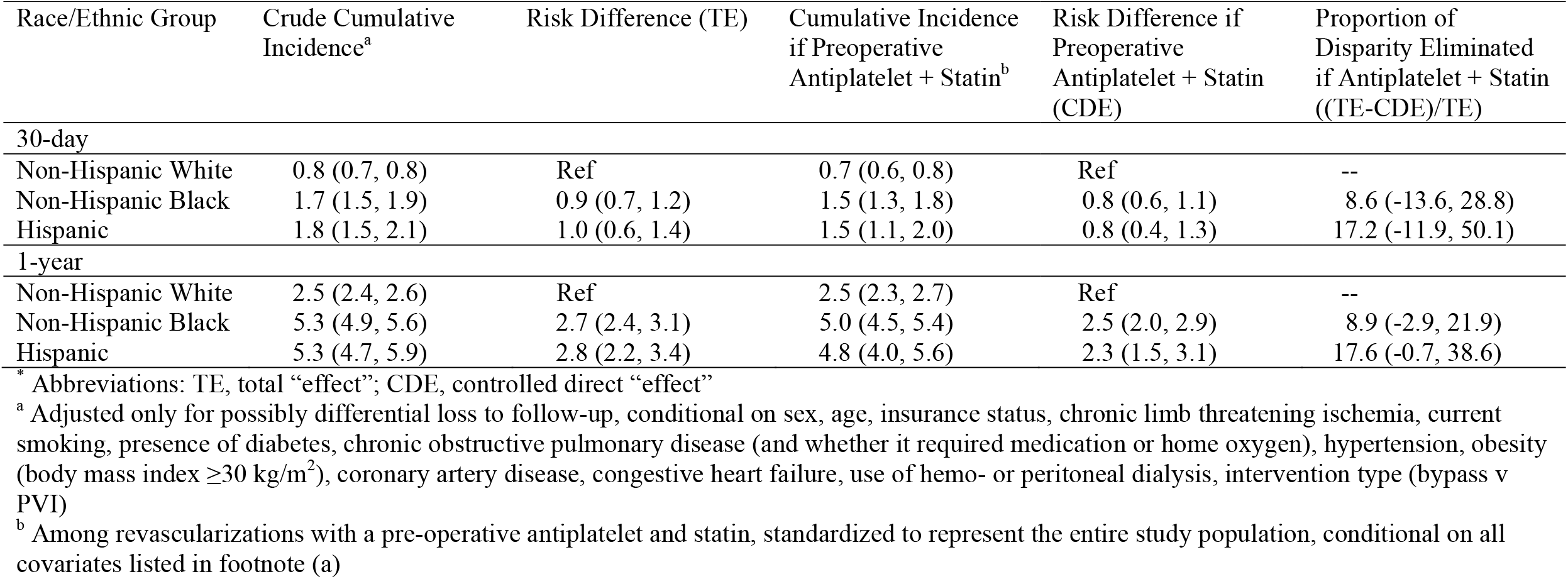
Cumulative incidence and racial/ethnic disparities of major amputations before and after a hypothetical intervention to ensure universal preoperative (within 36 hours) antiplatelet and statin for n=100,579 revascularizations for chronic limb-threatening ischemia or claudication, captured by the Vascular Quality Initiative registry, United States, 2011-2018

### Secondary Analyses, Revascularizations for CLTI

During our study, 55,451 limbs in 50,250 patients were treated with revascularization for CLTI. The 1-year risk of major amputation was 4.3% (95% CI: 4.1, 4.5) in non-Hispanic White patients, 7.6% (95% CI: 7.0, 8.1) in non-Hispanic Black patients, and 7.6% (95% CI: 6.8, 8.4) in Hispanic patients. We estimated that universal preoperative antiplatelet and statins would have reduced the disparity gap for non-Hispanic Black patients compared to non-Hispanic White patients by 7% (95% CI: -11,23) at one year; disparities in amputation risk for Hispanic patients compared to non-Hispanic White patients would have been reduced by 22% (95% CI: -1,46) at one year (Table V).

**Table V.**
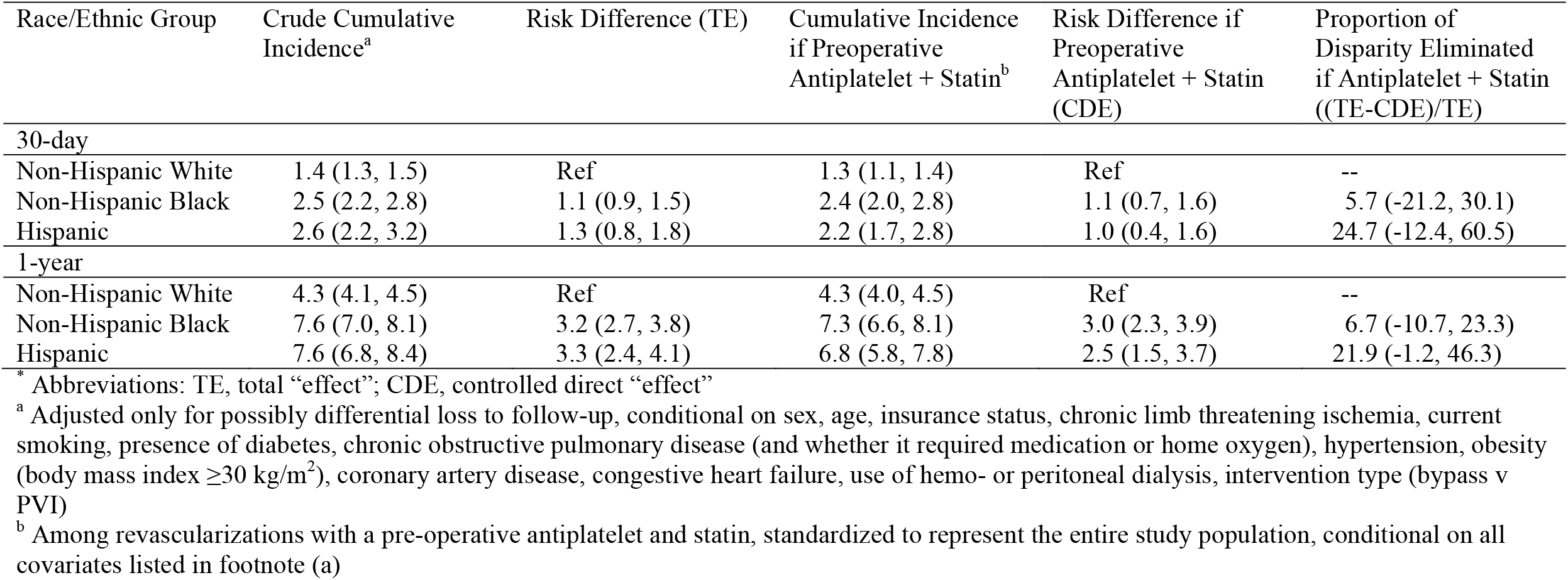
Cumulative incidence and racial/ethnic disparities of major amputations before and after a hypothetical intervention to ensure universal preoperative (within 36 hours) antiplatelet and statin for n=55,451 revascularizations for chronic limb-threatening ischemia, captured by the Vascular Quality Initiative registry, United States, 2011-2018

### Secondary Analyses, Revascularizations for Claudication

During our study, 45,128 limbs in 38,349 patients were treated with revascularization for claudication. The 1-year risk of amputation for non-Hispanic Black patients (1.1%; 95% CI: 0.8,1.4) was higher than for non-Hispanic White patients (0.6%; 95% CI: 0.5,0.6); the 1-year risk for Hispanic patients was 0.9% (95% CI: 0.5,1.3). We estimated that the proportion of racial/ethnic disparities eliminated by preoperative antiplatelet and statins would be 73% (95% CI: 39, 139) for non-Hispanic Black patients and by 47% (95% CI: -315, 502) for Hispanic patients (Table VI).

**Table VI.** Cumulative incidence and racial/ethnic disparities of major amputations before and after a hypothetical intervention to ensure universal preoperative (within 36 hours) antiplatelet and statin for n=45,128 revascularizations for claudication, captured by the Vascular Quality Initiative registry, United States, 2011-2018

| Race/Ethnic Group | Crude Cumulative Incidence<sup>a</sup> | Risk Difference (TE) | Cumulative Incidence if Preoperative Antiplatelet + Statin<sup>b</sup> | Risk Difference if Preoperative Antiplatelet + Statin (CDE) | Proportion of Disparity Eliminated if Antiplatelet + Statin ((TE-CDE)/TE) |
| --- | --- | --- | --- | --- | --- |
| 30-day |  |  |  |  |  |
| Non-Hispanic White | 0.1 (0.1, 0.2) | Ref | 0.1 (0.1, 0.1) | Ref | -- |
| Non-Hispanic Black | 0.2 (0.1, 0.4) | 0.1 (-0.0, 0.2) | 0.1 (0.0, 0.1) | -0.0 (-0.1, 0.0) | 152.9 (-289.7, 846.0) |
| Hispanic | 0.1 (0.0, 0.2) | -0.0 (-0.1, 0.2) | 0.1 (0.0, 0.4) | 0.0 (-0.1, 2.4) | 341.9 (-592.9, 1100.6) |
| 1-year |  |  |  |  |  |
| Non-Hispanic White | 0.6 (0.5, 0.6) | Ref | 0.5 (0.4, 0.6) | Ref | -- |
| Non-Hispanic Black | 1.1 (0.8, 1.4) | 0.5 (0.2, 0.8) | 0.7 (0.4, 1.0) | 0.1 (-0.1, 0.4) | 72.5 (38.7, 138.8) |
| Hispanic | 0.9 (0.5, 1.3) | 0.4 (-0.1, 0.8) | 0.7 (0.3, 1.3) | 0.2 (-0.3, 0.7) | 47.4 (-314.9, 502.4) |

\* Abbreviations: TE, total “effect”; CDE, controlled direct “effect”
<sup>a</sup> Adjusted only for possibly differential loss to follow-up, conditional on sex, age, insurance status, chronic limb threatening ischemia, current smoking, presence of diabetes, chronic obstructive pulmonary disease (and whether it required medication or home oxygen), hypertension, obesity (body mass index $\geq 30$ kg/m<sup>2</sup>), coronary artery disease, congestive heart failure, use of hemo- or peritoneal dialysis, intervention type (bypass v PVI)
<sup>b</sup> Among revascularizations with a pre-operative antiplatelet and statin, standardized to represent the entire study population, conditional on all covariates listed in footnote (a)

## DISCUSSION

Among patients treated with infrainguinal revascularization for symptomatic PAD, Black and Hispanic patients had a 1-year risk of amputation approximately two times the risk among non-Hispanic White patients. While this paper focused on potential ways to reduce those disparities, the observed race/ethnicity disparities represent actual human suffering and we hope this finding invigorates the vascular community to work to mitigate these outcomes. As one potential explanation for disparities in amputation risk, we investigated the role of sub-optimal prescription of preoperative antiplatelet and statins. We found that only 60% of patients had been prescribed preoperative antiplatelet and statin medications. We found only minimal differences in the prescription of these medications across race/ethnicity. However, we found the effect of antiplatelet and statins was greater in Black and Hispanic patients such that, had all patients received the appropriate guideline recommended medications, 1) risk of amputation would have been lower for all patients and 2) a significant portion of the disparities in outcomes experienced by Black and Hispanic patients relative to White patients would have been eliminated. As such, we have quantified an actionable way to reduce the major limb amputation disparities that have been documented for decades.

We estimated the largest proportion of disparities could be eliminated by properly administering antiplatelet and statin medications to patients with claudication, a supposedly non-limb threatening condition. While differences in the overall low rate of pre-operative antiplatelet and statins across our three race/ethnicity groups were minimal, Black and Hispanic patients did have a lower prevalence of antiplatelet and statin use. This finding of poor care aligns with literature on diseases such as lung cancer, where Black women have been shown to have less access to screening and early stage surgery^30^. First line therapy for claudication should be cardiovascular risk reduction with medical therapy and revascularization procedures should be reserved for those with very short distance claudication who have failed non-interventional management, including supervised exercise therapy to improve walking distances. More research is needed into why all patients, and especially Black and Hispanic patients, are not treated with best medical practices when they present for a revascularization procedure. It will also be important to study the circumstances that cause these individuals to be referred for vascular specialty care and to increase awareness of claudication in primary care settings where antiplatelet and statin could be initially prescribed.

To explain higher rates of amputation in Black patients, some studies have suggested that Black patients have a genetic predisposition to PAD and present at advanced stages with more anatomically complex disease^31, 32^. History tells us that the majority of the time that racial disparities in health outcomes are hypothesized to be due to “ genes,” better explanations rooted in systemic racism emerge. Worse, a quest for an explanation that blames the patient either indirectly (by faulting their genes) or directly (by suggesting they seek care late) ignores the role of systemic racism in determining whether they develop PAD (e.g., access to a healthy diet and safe spaces and leisure time for exercise, access to earlier medical therapy), when they present for care, how quickly their condition is diagnosed, what primary and secondary preventive care is available to them, and (the intervention of next-to-last-resort) the circumstances under which they receive vascular intervention for symptomatic PAD. Additionally, as in other conditions where pain is the primary symptom^33, 34^, it is possible that the severity of claudication is underestimated in Black and Hispanic patients and they are not referred until their claudication is more severe than in white patients. Although we describe a potential opportunity to reduce health disparities by increasing prescription and uptake of antiplatelet and statins among patients undergoing a revascularization procedure, limiting ourselves to interventions at such late stages of PAD care will always fall short of eliminating health disparities. However, the point at which patients present for vascular surgery represents an inflection point where we have an opportunity to, at the very least, not further propagate disparities.

There are other ACC/AHA Class I recommendations that define best practices for medical management, imaging methods to assess anatomy, and specific operative techniques. We found that provision of preoperative antiplatelet and statin may reduce amputation disparities. Optimized management of hypertension, diabetes, and smoking represent other critical areas of preventive care that should be explored. Assuming our estimate for the proportion of disparities reduced by universal uptake of preoperative antiplatelet/statin was correct, small incremental differences in each measure of optimized care may lead to the major disparities we see in outcomes.

Our results are limited to patients who underwent lower extremity revascularization for symptomatic PAD. There are selection processes that drive the patient population who were eligible for this study, for example, access to health care for diagnosis of PAD and initial revascularization surgery. These selection processes are associated with race and ethnicity, such that Black and Hispanic patients who undergo revascularization may be a healthier subset of all disadvantaged patients who would be eligible for this procedure, as compared to white patients. If those disparities were remedied early in the prevention and treatment pathways, we might hypothesize that the observed disparities after revascularization would be more dramatically reduced (even prior to additional interventions such as increasing adherence to guideline-based care).

The VQI registry was not designed specifically for research purposes and follow-up past one year is limited. Registry information is updated manually and is subject to human error. However, we report longer term outcomes (one year) than many previous publications. Additionally, the VQI is a “ real world” data source that provides a view of current outcomes in a large population of patients undergoing lower extremity revascularization procedures for symptomatic PAD in both tertiary and community hospitals. Thus, our results are likely more generalizable to PAD patients undergoing revascularization in the United States than prior reports.

There are a range of patient, physician, and health care system factors that impact the provision of cardiovascular care^35^. Our study examined one facet of this conceptual model - the prescription of appropriate medications by physicians that surgically manage PAD. We found that a substantial proportion of patients in this study did not receive appropriate medications prior to their revascularization attempt. We are not the first to find that uptake of optimal medical care is low among patients with PAD^36^ or that non-optimized medical care is connected to poor outcomes^20, 37^. In fact, Black and Hispanicpatients with PAD have had worse limb outcomes than white patients with PAD for decades and the studies that connected guideline concordant care to better outcomes are from ten years ago^19^. However, our study does not simply document this but instead identifies and quantifies an actionable way to reduce amputation disparities moving forward. We provide clear evidence that prescription of antiplatelet and statin, an ACC/AHA Class I recommendation, is a guideline that should be followed and improves outcomes for all, while closing outcome gaps for vascular patients from disadvantaged racial/ethnic groups. Finally, while our study documents the deficiencies in care and identified an actionable way to reduce disparities in outcomes, the mechanisms behind why guidelines are or are not followed remains an unanswered and critically important area of future study.

## Data Availability

In general, all VQI data is protected by the SVS PSO Data Analysis Guidelines for Use. Prospective investigators are required to submit a formal proposal to the National Research Advisory Committee (RAC). Investigators have access to data from any of the 14 modules to which their center has subscribed.

---

**Figure S1.**
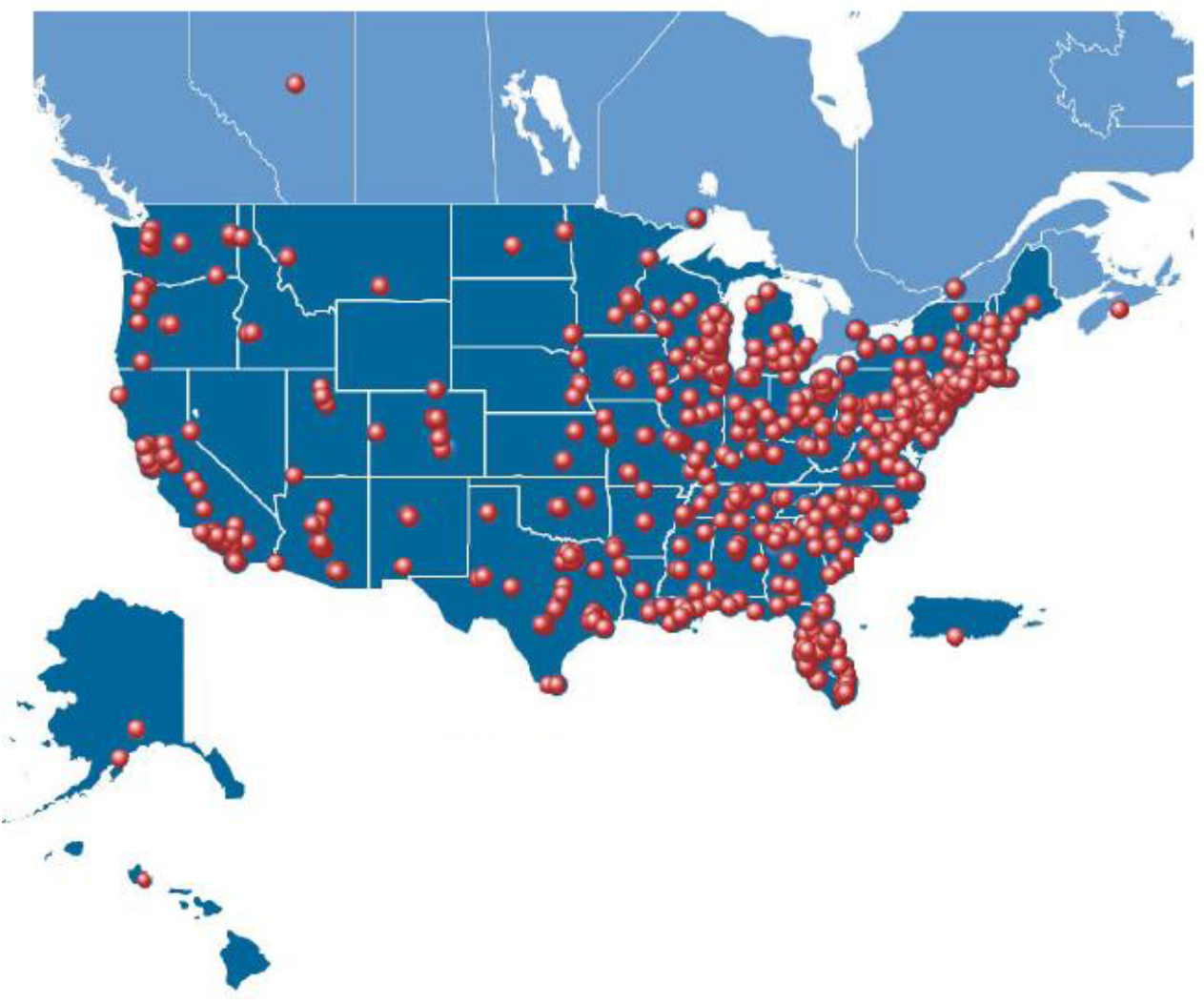
Map of Vascular Quality Initiative Sites.

## REFERENCES

1. Eid MA, Mehta KS, Goodney PP. Epidemiology of peripheral artery disease. Semin Vasc Surg. 2021;34:38–46

2. Fowkes FGR, Rudan D, Rudan I, Aboyans V, Denenberg JO, McDermott MM, et al. Comparison of global estimates of prevalence and risk factors for peripheral artery disease in 2000 and 2010: A systematic review and analysis. The Lancet. 2013;382:1329–1340

3. Virani SS, Alonso A, Benjamin EJ, Bittencourt MS, Callaway CW, Carson AP, et al. Heart disease and stroke statistics-2020 update: A report from the american heart association. Circulation. 2020;141:e139–e596

4. Allison MA, Ho E, Denenberg JO, Langer RD, Newman AB, Fabsitz RR, et al. Ethnic-specific prevalence of peripheral arterial disease in the united states. Am J Prev Med. 2007;32:328 –333

5. Chen L, Zhang D, Shi L, Kalbaugh CA. Disparities in peripheral artery disease hospitalizations identified among understudied race-ethnicity groups. Front Cardiovasc Med. 2021;8:692236

6. Kalbaugh C, Kucharska-Newton A, Wruck L, Lund JL, Selvin E, Matsushita K, et al. Peripheral artery disease prevalence and incidence estimated from both outpatient and inpatient settings among medicare fee-for-service beneficiaries in the atherosclerosis risk in communities (aric) study. JAHA. 2017;(in press)

7. Kalbaugh CA, Loehr L, Wruck L, Lund JL, Matsushita K, Bengtson LG, et al. Frequency of care and mortality following an incident diagnosis of peripheral artery disease in the inpatient or outpatient setting: The aric (atherosclerosis risk in communities) study. Journal of the American Heart Association. 2018;7:e007332

8. Arya S, Binney Z, Khakharia A, Brewster LP, Goodney P, Patzer R, et al. Race and socioeconomic status independently affect risk of major amputation in peripheral artery disease. J Am Heart Assoc. 2018;7

9. Holman KH, Henke PK, Dimick JB, Birkmeyer JD. Racial disparities in the use of revascularization before leg amputation in medicare patients. J Vasc Surg. 2011;54:420–426

10. Durazzo TS, Frencher S, Gusberg R. Influence of race on the management of lower extremity ischemia: Revascularization vs amputation. JAMA Surg. 2013;148:617–623

11. Goodney PP, Travis LL, Nallamothu BK, Holman K, Suckow B, Henke PK, et al. Variation in the use of lower extremity vascular procedures for critical limb ischemia. Circulation: Cardiovascular Quality and Outcomes. 2012;5:94–102

12. Criqui MH, Aboyans V. Epidemiology of peripheral artery disease. Circ Res. 2015;116:1509–1526

13. Allison MA, Ho E, Denenberg JO, Langer RD, Newman AB, Fabsitz RR, et al. Ethnic-specific prevalence of peripheral arterial disease in the united states. American journal of preventive medicine. 2007;32:328–333

14. Hua S, Isasi CR, Kizer JR, Matsushita K, Allison MA, Tarraf W, et al. Underuse of cardiovascular medications in individuals with known lower extremity peripheral artery disease: Hchs/sol. J Am Heart Assoc. 2020;9:e015451

15. Mustapha J, Fisher BT, Rizzo JA, Chen J, Martinsen BJ, Kotlarz H, et al. Explaining racial disparities in amputation rates for the treatment of peripheral artery disease (pad) using decomposition methods. Journal of racial and ethnic health disparities. 2017;4:784–795

16. Hughes K, Seetahal S, Oyetunji T, Rose D, Greene W, Chang D, et al. Racial/ethnic disparities in amputation and revascularization: A nationwide inpatient sample study. Vascular and endovascular surgery. 2014;48:34–37

17. Gerhard-Herman M, Gornik H, Barrett C, Barshes N, Corriere M, Drachman D, et al. 2016 aha/acc guideline on the management of patients with lower extremity peripheral artery disease: Executive summary: A report of the american college of cardiology/american heart association task force on clinical practice guidelines.. Journal of the American College of Cardiology. 2017;69:1465–1508

18. Hirsch AT, Haskal ZJ, Hertzer NR, Bakal CW, Creager MA, Halperin JL, et al. Acc/aha 2005 guidelines for the management of patients with peripheral arterial disease (lower extremity, renal, mesenteric, and abdominal aortic): A collaborative report from the american association for vascular surgery/society for vascular surgery,* society for cardiovascular angiography and interventions, society for vascular medicine and biology, society of interventional radiology, and the acc/aha task force on practice guidelines (writing committee to develop guidelines for the management of patients with peripheral arterial disease). Journal of the American College of Cardiology. 2006;47:e1–e192

19. Armstrong EJ, Chen DC, Westin GG, Singh S, McCoach CE, Bang H, et al. Adherence to guideline-recommended therapy is associated with decreased major adverse cardiovascular events and major adverse limb events among patients with peripheral arterial disease. Journal of the American Heart Association. 2014;3:e000697

20. Vogel TR, Dombrovskiy VY, Galinanes EL, Kruse RL. Preoperative statins and limb salvage after lower extremity revascularization in the medicare population. Circ Cardiovasc Interv. 2013;6:694–700

21. Hussain MA, Al-Omran M, Mamdani M, Eisenberg N, Premji A, Saldanha L, et al. Efficacy of a guideline-recommended risk-reduction program to improve cardiovascular and limb outcomes in patients with peripheral arterial disease. JAMA Surgery. 2016;151

22. Cronenwett JL, Kraiss LW, Cambria RP. The society for vascular surgery vascular quality initiative. J Vasc Surg. 2012;55:1529–1537

23. MacCallum K, Yau P, Phair J, Lipsitz E, Scher L, Garg K. Ambulatory status following major lower extremity amputation.. Annals of vascular surgery. 2021;Feb 1;71:331–337

24. Govsyeyev N, Nehler MR, Low Wang CC, Kavanagh S, Hiatt WR, Long C, et al. Etiology and outcomes of amputation in patients with peripheral artery disease: Insights from the euclid trial. J Vasc Surg. 2021

25. Long CA, Mulder H, Fowkes FGR, Baumgartner I, Berger JS, Katona BG, et al. Incidence and factors associated with major amputation in patients with peripheral artery disease: Insights from the euclid trial. Circ Cardiovasc Qual Outcomes. 2020;13:e006399

26. Heart TCCotNY, Association. Diseases of the heart and blood vessels. Boston: Little, Brown & Co. ; 1995.

27. Cole SR, Lau B, Eron JJ, Brookhart MA, Kitahata MM, Martin JN, et al. Estimation of the standardized risk difference and ratio in a competing risks framework: Application to injection drug use and progression to aids after initiation of antiretroviral therapy. Am J Epidemiol. 2015;181:238–245

28. Robins JM, Finkelstein DM. Correcting for noncompliance and dependent censoring in an aids clinical trial with inverse probability of censoring weighted (ipcw) log-rank tests. Biometrics. 2000;56:779–788

29. Howe CJ, Dulin-Keita A, Cole SR, Hogan JW, Lau B, Moore RD, et al. Evaluating the population impact on racial/ethnic disparities in hiv in adulthood of intervening on specific targets: A conceptual and methodological framework. Am J Epidemiol. 2018;187:316–325

30. Cykert S, Dilworth-Anderson P, Monroe MH, Walker P, McGuire FR, Corbie-Smith G, et al. Factors associated with decisions to undergo surgery among patients with newly diagnosed early-stage lung cancer. Jama. 2010;303:2368–2376

31. Rucker-Whitaker C, Feinglass J, Pearce WH. Explaining racial variation in lower extremity amputation. Arch Surg. 2003;138:1347–1351

32. Rivero M, Nader ND, Blochle R, Harris LM, Dryjski ML, Dosluoglu HH. Poorer limb salvage in african american men with chronic limb ischemia is due to advanced clinical stage and higher anatomic complexity at presentation. Journal of vascular surgery. 2016;63:1318–1324

33. Befus DR, Irby MB, Coeytaux RR, Penzien DB. A critical exploration of migraine as a health disparity: The imperative of an equity-oriented, intersectional approach. Curr Pain Headache Rep. 2018;22:79

34. Madden EF, Qeadan F. Racial inequities in u.S. Naloxone prescriptions. Substance Abuse. 2020;41:232–244

35. Kressin N, Peterson L. Racial differences in the use of invasive cardiovascular procedures: Review of the literature and prescription for future research. Ann Intern Med. 2001;135:352–366

36. Saxon JT, Safley DM, Mena-Hurtado C, Heyligers J, Fitridge R, Shishehbor M, et al. Adherence to guideline-recommended therapy-including supervised exercise therapy referral-across peripheral artery disease specialty clinics: Insights from the international portrait registry. J Am Heart Assoc. 2020;9:e012541

37. Armstrong EJ, Wu J, Singh GD, Dawson DL, Pevec WC, Amsterdam EA, et al. Smoking cessation is associated with decreased mortality and improved amputation-free survival among patients with symptomatic peripheral artery disease. J Vasc Surg. 2014;60:1565–1571

